# Peri Operative deLta rEnin ConcentrATion (POLECAT) Study Protocol and Analysis Plan

**DOI:** 10.64898/2026.05.26.26352884

**Authors:** Naomi Boyer, Syeda Haider, Charles Piercy, Alexander Zarbock, Theophilus Luke Samuels, Aikaterini Papadopoulou, Lui G. Forni, Ben Creagh-Brown

## Abstract

**Background:** Post-operative hypotension and vasoplegia are well recognised following cardiac surgery but remain poorly characterised after major non-cardiac surgery, despite associations with acute kidney injury (AKI), cardiovascular complications, and increased mortality. Dysregulation of the renin–angiotensin–aldosterone system (RAAS) may underpin haemodynamic instability in this setting, yet data in abdominal surgery are limited.

**Objectives:** The POLECAT (Perioperative delta Renin) study aims to determine whether changes in circulating renin concentration (Δrenin) from pre-operative baseline to the early post-operative period are associated with post-operative vasoplegia in patients undergoing major abdominal surgery requiring intensive care admission.

**Methods:** POLECAT is a single-centre, prospective observational study conducted at a UK tertiary referral hospital. Adult patients undergoing planned or emergency abdominopelvic surgery with anticipated intensive care admission are enrolled. Blood samples are obtained pre-operatively, within four hours post-operatively, and on post-operative day one to measure renin and a panel of endothelial, renal, and immune biomarkers. The primary outcome is post-operative vasoplegia, defined as the requirement for a vasopressor infusion at 08:00 on post-operative day one. Secondary outcomes include alternative vasoplegia definitions, AKI (KDIGO criteria), vasopressor burden, organ dysfunction, cardiovascular complications, length of stay, and mortality. Multivariable regression, receiver operating characteristic analyses, and predefined subgroup analyses will be performed, with sensitivity analyses addressing missing data.

**Conclusions:** This study will clarify the relationship between peri-operative RAAS dysfunction and vasoplegia following major abdominal surgery. Findings may support biomarker-guided risk stratification and inform future interventional trials targeting haemodynamic instability in this high-risk population.

## Background

### Rationale

Post operative hypotension and vasoplegia following cardiac surgery have been extensively explored within literature, and is associated with complications, however this phenomenon is less commonly reported following non-cardiac surgery. It is known that hypotension following non cardiac surgery is associated with multiple adverse events including increases in post operative acute kidney injury (AKI), myocardial infarction, cerebrovascular injury and 30 and 90 day mortality [1–4]. Previous literature has found that 19% of gynae-oncology patients exhibit post operative vasoplegia [5]. Given that non-cardiac surgery is more commonly performed the minority of patients that develop complications account for the overwhelming majority of associated morbidity [6] downstream healthcare interventions and resource use, substantially impacting the efficiency of an overburdened healthcare system and impacting on the care available to other patients [7].

### Literature

The renin angiotensin aldosterone system (RAAS) plays a key role in maintaining haemodynamic stability, vascular tone and electrolyte homeostasis. It has increasingly been recognized as being dysfunctional [8] during critical illness.

Surgery causes alterations in the RAAS-axis, typically with increases in renin after the operation. The usual relationship between increased renin and increased angiotensin II resulting in an increased blood pressure is diminished post operatively due to i) reduced ACE activity (usually converts Angiotensin I to II), ii) reduced expression of the receptor for Angiotensin II [7]. It has been demonstrated that in cardiac surgery, those with the greatest renin increment (delta renin, Δrenin) pre to post operatively had a higher incidence of AKI, lower blood pressure and longer duration of vasopressor therapy [8].

Multiple randomized controlled trials [9–11] are now investigating renin-guided angiotensin II therapy in both cardiac surgery and septic shock. High baseline renin levels (indicating failure of RAAS homeostasis) have been associated with improved response to angiotensin II therapy [9].

### Knowledge gaps

Biomarkers offer an attractive prospect for the identification of those with a propensity to develop post operative vasoplegia as well as those who will develop more profound post operative complications.

Given it is unknown whether the post operative hypotension that is exhibited following abdominal surgery is a pathophysiologically distinct entity from that exhibited in other aetiologies such as post cardiac surgery or sepsis, biomarkers can also offer a potential insight into the mechanisms implicated within development of vasoplegia following abdominal surgery.

Biomarkers have been investigated within the literature, although none within non cardiac surgery [12]. A recent systematic review identified significant gaps within the literature both regarding a consensus definition of vasoplegia and with the utilisation of biomarkers [12].

## Study Objectives

### Primary objective

To assess whether there is an association between higher Δrenin (change in circulating renin concentration from pre-operative to 4 hours post operative levels) and post operative vasoplegia in patients undergoing major abdominal surgery requiring intensive care unit admission.

### Secondary objectives

1. To assess whether there is an associated between higher Δrenin and different post operative vasoplegia definitions (specified below).
2. To determine the overall burden of vasopressor use post operatively in this surgical cohort
3. To determine the incidence of post operative AKI
4. To characterize temporal trends of renin concentrations post operatively following major abdominal surgery
5. To assess associations between changes in renin and development of AKI
6. To examine associations between type of surgery and incidence of post operative hemodynamic instability and renin measurements, with specific emphasis on patients undergoing emergency surgery with peritonitis
7. To assess associations between regular medications that alter the renin-angiotensin-aldosterone system (RAAS) and changes in renin concentrations
8. To examine associations between endothelial biomarkers (Bio-ADM syndecan-1) and post operative vasoplegia, AKI, and organ dysfunction
9. To assess the performance of proenkephalin (penKid) as an early biomarker of post operative AKI

### Exploratory objectives

1. To examine associations between immune activation biomarkers (circulating DPP3, DPP4, suPAR) and post operative vasoplegia, organ dysfunction, and clinical outcomes
2. To identify distinct pathophysiological subphenotypes based on angiopoietin profiles (Angiopoietin-1/Angiopoietin-2 ratio)
3. To identify inflammatory subphenotypes using cytokine profiles and latent class analysis
4. To explore mechanistic relationships between RAAS dysfunction, endothelial injury, immune dysregulation, and adverse post operative outcomes
5. To generate hypotheses regarding potential therapeutic targets for future interventional trials

## Methods

### Study Design

This is an observational study, to be performed at the Royal Surrey County Hospital, which is a 450-bed tertiary centre for upper-gastrointestinal, gynae-oncology, hepatobiliary and urology surgery (referral centre for >1million patients); both planned and unplanned emergency general surgical procedures. Recruitment took place between November 2023 and June 2025. All surgical patients who have an intensive care bed booked, will be screened by members of the research team and research fellows. The blood sampling required will be taken from invasive lines that are routinely inserted during the surgical procedures being investigated, no further venepuncture is required. If enrolled into the study they will have baseline blood taken at the time of arterial line or central line insertion, prior to knife to skin. A second sample will again be taken on arrival to the ICU where it is standard protocol to take a full set of blood investigations including blood gases. The final samples will be acquired day 1 post operatively. See Figure 1.0 for schedule of assessments.

**Figure 1:**
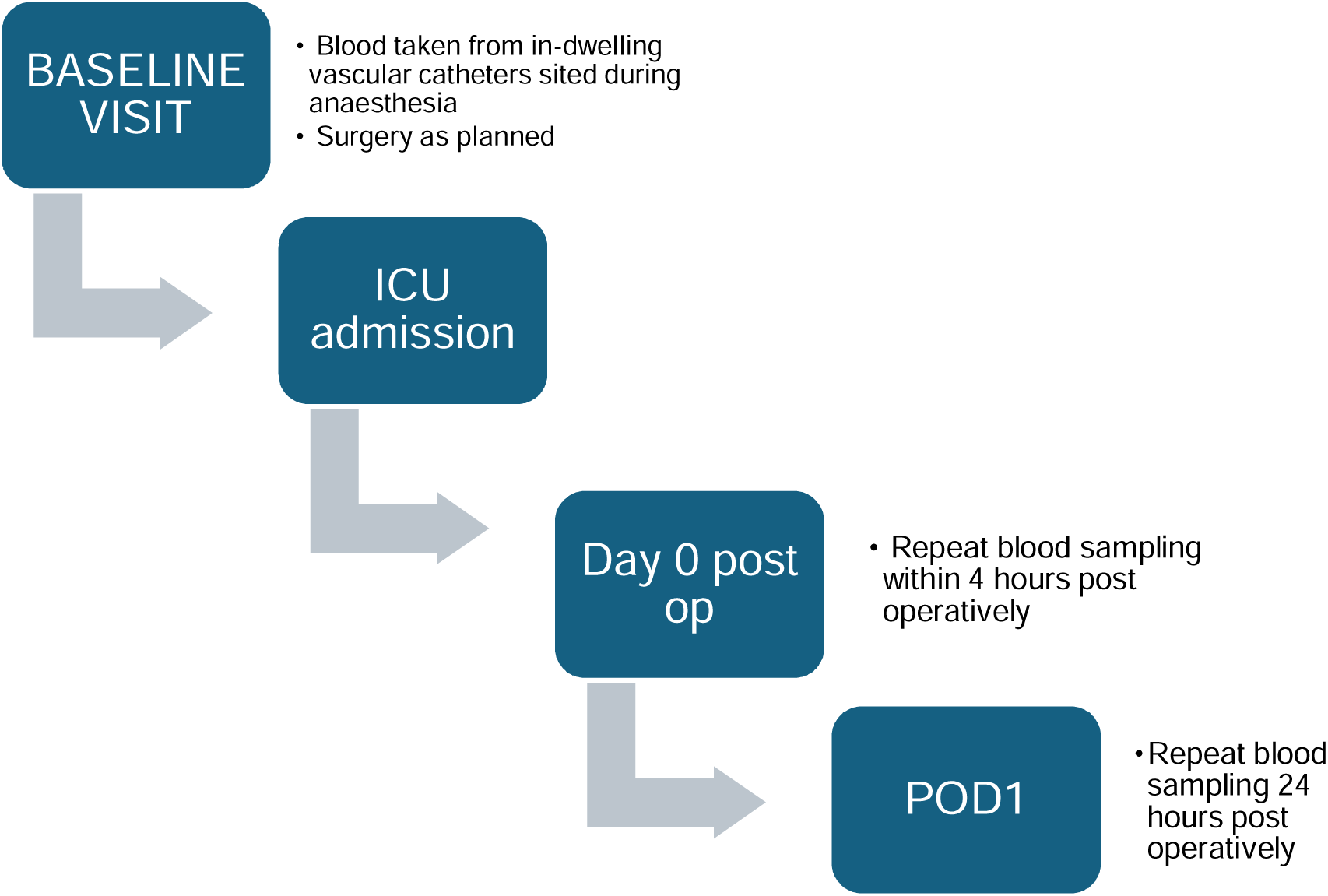
Schedule of assessments.

Data collection will include the quantity of intravenous fluid administered intra-operatively as well as in the first 72 hours of ICU admission, burden of vasopressor usage, urine output and urea and creatinine changes from baseline and over the first 72 hours of ICU admission. This will be obtained from electronic notes.

### Participants

#### Inclusion Criteria

- Age ≥18 years
- All patients having abdominopelvic surgery (planned or unplanned) needing ICU admission where arterial line or central line has been placed
- Able to provide consent, or declaration of agreement from personal or professional consultee

#### Exclusion Criteria

- Patient aged < 18 years
- Patients already receiving vasopressor support pre-operatively
- Patients receiving palliative care at the time of recruitment or expected to die within next 24 hours
- Patients receiving recent dialysis or with end stage kidney disease
- Patient or consultee unable to communicate in verbal and written English
- Patients held in an institution by legal or official order
- Patients with severe mental health disorders that might impair their capacity
- Adrenal surgery, such as resection for phaeochromocytoma

The exclusion criteria will be confirmed through patient’s medical records or by asking the patient.

#### Minimising bias

Selection bias has been minimized through systematic screening of all eligible surgical cases. Information bias has been reduced through standardized data collection procedures, validated eCRF with range checks, and source document verification. The primary outcome (vasoplegia at 08:00 on post operative day 1 (POD1)) is objectively defined and time-stamped, reducing measurement bias. For exploratory biomarker analyses, laboratory personnel are blinded to clinical outcomes. Loss to follow-up is minimized by using hospital records and national death registry; sensitivity analyses will assess impact of missing outcome data.

#### Variables

##### Exposures

- Primary: Δrenin (pre-operative to 4hours post operative)
- Secondary biological exposures: Bio-ADM, SDC-1, penKid, cDPP3, DPP4, suPAR, Angiopoietins, sTNFR-1

##### Outcomes

- Primary: Post operative vasoplegia
- Secondary: alternative vasoplegia definitions, AKI, vasopressor duration, mechanical ventilation, ICU/hospital LOS, cardiovascular complications, mortality

##### Predictors/Confounders

- Patient factors: Age, sex, weight, frailty, comorbidities
- Surgical factors: Surgery type, urgency, approach, duration
- Anaesthetic factors: Epidural use, intra-operative vasopressors, fluid balance
- Medications: RAAS-modifying drugs and perioperative management
- Baseline labs: Creatinine, haemoglobin

##### Effect Modifiers (Planned Subgroup Analyses)

- Emergency vs elective surgery
- Presence of peritonitis
- Use of RAAS-modifying medications
- Baseline renal function
- Thoracic epidural use

This comprehensive variable definition and data source documentation ensures methodological transparency, facilitates data interpretation, and supports reproducibility of the POLECAT study findings. All variables are recorded in the hospital electronic patient record (Oracle Millennium) apart from the experimental assay results.

#### Sample size calculation

Using logistic regression and assuming a conservative estimate of 15% incidence of vasoplegia (based off knowledge that previous work that has demonstrated 19% of gynae-oncology patients in our centre have vasoplegia post operatively). Previous work has shown that patients with a high Δrenin had 3 times the incidence of vasoplegia compared to the patients with low Δrenin, we can assume that the incidence of vasoplegia will be 5% in our patients without high Δrenin With an alpha of 0.05 and power of 0.8 we need a sample size of 106 per group, making a total of 212. Allowing for a 35% loss, this means we need 285 patients.

### Data Collection

#### Patient Identification and Consent

Daily screening of surgical schedules identifies eligible patients undergoing abdominopelvic surgery requiring ICU admission. Written informed consent is obtained pre-operatively for elective cases. For emergency surgery patients lacking capacity, consultee agreement is sought with prospective consent obtained when capacity regained.

#### Baseline Data Collection

Pre-operative assessment includes demographics (age, sex, weight, height), Clinical Frailty Score, comorbidities with sub-classifications, comprehensive medication history (emphasizing RAAS-modifying agents and perioperative management), ASA classification, and baseline laboratory values (creatinine, haemoglobin, albumin). Surgical details include indication, planned procedure, and urgency. Data are abstracted from electronic health records and documented in the electronic case report form (eCRF).

#### Blood Sample Collection and Processing

Three timepoints: (1) Pre-operative at arterial/central line insertion, (2) Within 4 hours of ICU admission, (3) Morning of post operative day 1 (POD1) with routine labs.

Procedure: 10mL collected via EDTA tube from existing vascular access. Samples centrifuged within 15 minutes (2000-3000 rpm, 10 minutes, 2-8°C), plasma separated and aliquoted, then stored at −80°C with documented chain of custody.

Biomarkers by timepoint

- Pre-operative and 4h post-op: Renin, Bio-ADM, angiopoietin-1/2, sTNFR-1, cDPP3, DPP4, suPAR, syndecan-1
- 4h post-op only: Proenkephalin (penKid)
- Post operative day 1: Renin, Bio-ADM, cDPP3

Renin samples will be shipped on dry ice to University Hospital Münster for batch analysis. Bio-ADM samples will be shipped and analysed by Sphingotech. Other biomarkers analyzed via ELISA or automated immunoassays with quality control including duplicate assays and blinded laboratory personnel

#### Intra-operative Data Collection

Anaesthetic records provide surgery duration, epidural use, vasopressor administration (bolus and infusion details including agent, concentration, maximum rate), crystalloid volume, blood products, intra-operative urine output, and estimated blood loss. Data abstracted from electronic anaesthetic records and smart pump systems.

#### Post operative Monitoring

Primary outcome assessment (08:00 on POD1): Precise documentation of vasopressor status, including presence/absence of infusions, specific agents and rates, patient weight, and epidural status. Two-person verification with time-stamped documentation ensures quality. Additional daily data (Days 0-7) will be collected including: maximum vasopressor doses for each agent, mechanical ventilation status, daily creatinine and urine output, fluid balance, SOFA scores, and complications including cardiovascular events (MI, new atrial fibrillation), infections requiring antibiotics, and surgical complications (Accordion classification [13]). Cumulative vasopressor duration calculated from timestamped infusion records.

#### Outcomes and Follow-up

ICU and hospital length of stay recorded from admission/discharge timestamps. Discharge destinations documented. Thirty-day vital status determined via electronic health record review, telephone contact using standardized script, or national death registry check. Location and health status documented for survivors; date and location of death recorded for decedents.

#### Data Management and Quality Assurance

Electronic case report forms include automated validation with range checks, logic checks, and mandatory fields. Research staff complete standardized training with competency assessment. Query management system tracks discrepancies requiring resolution. Data stored on secure hospital servers with role-based access control, automated backups, and audit trails. Patient identifiers minimized; unique study IDs used with separate storage of linking log. All data and samples retained for minimum 15 years post-study completion.

### Flow Diagram of Study

### Ethical considerations

Ethical Approval and Regulatory Compliance

The study will be conducted according to the Declaration of Helsinki, ICH-GCP guidelines, and UK Policy Framework for Health and Social Care Research. Protocol approval has been obtained from a UK Research Ethics Committee prior to commencement (Wales research ethics committee + HRA reference: 23/WA/0150). Royal Surrey NHS Foundation Trust serves as sponsor.

#### Consent Procedures

Elective surgery patients: Written informed consent obtained pre-operatively after providing a Participant Information Sheet and allowing adequate time for consideration and questions.

Emergency surgery patients: For patients lacking capacity, advice is sought from a personal consultee (family member) or, if unavailable, a professional consultee (independent senior clinician) following Mental Capacity Act 2005 guidance. Retrospective consent is obtained when patients regain capacity post operatively.

Withdrawal: Participants may withdraw at any time without affecting clinical care, with options to retain or destroy previously collected data and samples.

#### Data Protection and Confidentiality

All data handling complies with UK GDPR and Data Protection Act 2018. Participants are assigned unique study IDs; a separate linking log to patient identifiers is stored securely with restricted access. Analysis datasets are fully de-identified with dates converted to relative timepoints. Electronic data are stored on secure NHS servers with role-based access controls, encryption, automated backups, and audit trails. Physical documents and consent forms are stored in locked cabinets in secure research offices. Biological samples are labelled only with study ID and visit number and stored in locked −80°C freezers with restricted access. For international biomarker analysis (University Hospital Münster, and Sphingotech), only de-identified samples and minimal clinical data are transferred using appropriate safeguards (Standard Contractual Clauses). Data and samples will be retained for minimum 15 years post-study completion per regulatory requirements.

### Data Management

Study data will be captured and managed using an electronic case report form (eCRF), with appropriate controls in place to ensure data integrity and confidentiality. Any physical records will be stored securely, and biological samples (where applicable) will be handled and stored in accordance with approved procedures. All personal data will be pseudonymised to protect participant identity.

Source documents will be clearly defined and maintained, with full traceability between source data and entries in the eCRF to support audit and verification requirements.

Study records will be retained for a minimum period of 15 years following study completion, after which data will be disposed of in line with documented disposal procedures and applicable regulatory requirements.

Oversight of the study will be provided by the Sponsor, supported by independent monitoring to ensure compliance with the protocol, regulatory standards, and good clinical practice.

### Study Governance

The study will be sponsored by Royal Surrey NHS Foundation Trust, which will assume overall responsibility for the initiation, management, and oversight of the research in accordance with applicable regulatory and governance requirements. Funding for this study is provided by the European Society of Intensive Care Medicine (ESICM).

Any potential conflicts of interest will be transparently disclosed. In particular, Professor Zarbock’s relevant relationships have been declared, and appropriate procedures are in place to identify, manage, and mitigate any actual or perceived conflicts in line with institutional and sponsor policies.

Overall responsibility for the day-to-day conduct of the study will rest with the Principal Investigator, who will ensure that the study is delivered in accordance with the approved protocol, ethical approvals, and applicable regulatory standards.

### Dissemination

Study results will be disseminated through publication in peer-reviewed journals and presentation at national and international conferences. Lay summaries will also be produced to support wider accessibility. The study protocol will be made available as a preprint to promote transparency.

Authorship will be determined in accordance with International Committee of Medical Journal Editors (ICMJE) criteria, with authorship order reflecting the nature and extent of individual contributions.

Individual participant data (IPD) will be made available upon reasonable request, subject to appropriate approvals and data protection requirements. The study protocol and statistical analysis plan will be publicly accessible, and sharing of biological samples may be considered in line with ethical and governance approvals.

## Statistical Analysis Plan

### Outcome Definitions

#### Primary Outcome: Post operative Vasoplegia

##### Definition 1 (Primary)

- Any patient receiving a vasopressor infusion at 08:00 hours on post operative day 1 (POD1), at any infusion rate.

##### Definition 2 (Secondary)

- Any patients receiving a vasopressor infusion where the noradrenaline dose (or equivalent) ≥0.1 mcg/kg/min at 08:00 hours on POD1 for all patients.

##### Definition 3 (Secondary)

This definition attempts to account for thoracic epidural vasodilatory effects and has tried to take a pragmatic approach based of clinical experience of the investigators

- For those patients without a thoracic epidural any dose vasopressor infusion at 08:00 hours on POD1 will be classed as vasopelgic
- For those patients with a thoracic epidural infusion running a vasopressor infusion where the noradrenaline dose or equivalent is ≥0.1 mcg/kg/min is required to be classed as vasoplegic

#### Secondary Outcomes

##### Acute Kidney Injury (AKI)

This will be assessed using KDIGO criteria based on serum creatinine highest severity will be reported and staged as KDIGO 1, 2, or 3. This will be assessed in the first 7 days post operatively [14].

##### Vasopressor Burden

Vasopressor burden will be assessed using the peak noradrenaline-equivalent dose (mcg/kg/min) within the first 72 hours post operatively, alongside cumulative vasopressor exposure measured as total hours of use within the first 7 days. Correlation analyses will examine relationships between peak dose, duration of exposure, and cumulative dose.

##### Organ Dysfunction Composite

Multi-organ dysfunction defined using daily SOFA scores with increase ≥2 points from baseline in any organ system, or composite of cardiovascular, renal, and respiratory dysfunction.

##### Cardiovascular Complications

Cardiovascular complications will include acute myocardial infarction (defined according to the Fourth Universal Definition [15]), new-onset atrial fibrillation (ECG confirmation), and other clinically significant dysrhythmias requiring therapeutic intervention. All cardiovascular outcomes will be assessed within the first 7 days following surgery.

##### Other Clinical Outcomes

- Mechanical ventilation requirement (first 7 days)
- Surgical complications (Accordion Classification grades 1-5)[13]
- Clinical suspicion for sepsis (first 7 days)
- ICU length of stay (days)
- Hospital length of stay (days)
- 30-day mortality

### Handling of Missing Data

#### Approach

Missing data will be reported descriptively including frequency and patterns. Reasons for missing data will be documented (not collected, test not performed, patient withdrawal, technical failure).

#### Primary Outcome (Vasoplegia)

Missing vasoplegia status at 08:00 POD1 will be treated as missing; no imputation will be performed for the primary outcome. Sensitivity analyses will explore impact of excluding patients with missing primary outcome data.

#### Biomarker Data

Missing baseline or post operative biomarker samples preclude calculation of change scores (Δrenin). These patients will be excluded from primary analysis but included in analyses using single timepoint values (e.g., baseline renin predicting outcomes).

#### Analysis Populations

Per protocol analysis will be performed with complete data for primary outcome (vasoplegia status at 08:00 POD1) and both pre-operative and 4-hour post operative renin measurements. Used for sensitivity analyses.

Intention-to-Treat analysis will be performed for all enrolled patients who provided consent and have at least one blood sample collected to enable calculation of single timepoint values.

#### Clinical Variables

For regression analyses, the primary approach will be complete case analysis. Where more than 10% of data are missing for key covariates, sensitivity analyses will be undertaken using multiple imputation. Multiple imputation by chained equations (MICE) will be applied, with 20 imputed datasets, with results combined using Rubin’s rules.

Patterns of missingness will be examined to assess whether data are Missing Completely at Random (MCAR), Missing at Random (MAR), or Missing Not at Random (MNAR), and findings will inform interpretation of the results.

#### Follow-up Data

Patients lost to 30-day follow-up will be censored at last known contact date for survival analyses. Vital status will be verified through hospital records and national death registry where available.

### Statistical Methods

#### General Principles

All analyses will be two-tailed with significance level α=0.05 unless otherwise specified. No adjustment for multiple comparisons will be made for secondary and exploratory analyses; these should be interpreted as hypothesis-generating. Continuous variables will be assessed for normality using Shapiro-Wilk test and visual inspection (histograms, Q-Q plots). Appropriate parametric or non-parametric tests will be selected based on distributional assumptions.

#### Descriptive Statistics

Continuous variables will be assessed for distributional assumptions using visual inspection and formal testing where appropriate. Normally distributed variables will be summarised as mean (standard deviation), and non-normally distributed variables as median (interquartile range). Minimum and maximum values will be reported where informative. Categorical variables will be summarised as counts and percentages, with cross-tabulation by outcome group where relevant.

Baseline characteristics will be presented in a descriptive table stratified by the primary outcome (vasoplegia versus no vasoplegia), including demographics, comorbidities, surgical variables, and baseline medication use. As the study is observational, no statistical testing for baseline group differences will be undertaken.

Biomarker concentrations will be summarised at each predefined timepoint using appropriate descriptive statistics based on distribution. Change scores (Δ values) will be calculated as post operative minus pre-operative concentrations and summarised accordingly.

All analyses and reporting will be conducted in accordance with the STROBE (Strengthening the Reporting of Observational Studies in Epidemiology) guidelines to ensure transparency, completeness, and reproducibility [16].

### Primary Analysis

#### Hypothesis

Patients with greater increment in circulating renin (Δrenin = renin at 4h post-op minus pre-op) will have higher incidence of vasoplegia (Definition 1).

#### Analytical approach

First, the ability of Δrenin to predict vasoplegia will be evaluated using receiver operating characteristic (ROC) curve analysis. The area under the curve (AUC) with 95% confidence intervals will be calculated to assess discriminatory performance. An optimal threshold for Δrenin will be identified using Youden’s Index.

Using this threshold, patients will be categorised into high versus low Δrenin groups. The incidence of vasoplegia between groups will be compared using a chi-squared test (or Fisher’s exact test where appropriate). Effect estimates will be reported as odds ratios with 95% confidence intervals, alongside diagnostic performance metrics including sensitivity, specificity, positive predictive value, and negative predictive value.

As a complementary analysis, Δrenin will also be analysed as a continuous variable. Mean or median Δrenin values will be compared between patients with and without vasoplegia using an independent t-test or Mann–Whitney U test, depending on distribution. Effect sizes will be reported to quantify the magnitude of association.

#### Reporting

Primary analysis results will include point estimates, 95% confidence intervals, p-values, and effect sizes. A CONSORT-style flow diagram will show patient enrolment, exclusions, and analysis populations.

### Secondary Analyses

#### Alternative vasoplegia definitions

The primary analysis will be repeated using Definitions 2 and 3 of vasoplegia. Consistency of associations between Δrenin and vasoplegia across definitions will be examined to assess robustness of findings.

##### Biomarker trajectories over time

Changes in biomarker concentrations across timepoints (pre-operative, 4 hours post operative, and post operative day 1) will be compared between patients with and without vasoplegia. A mixed-design ANOVA will be used for normally distributed data, and a non-parametric alternative with appropriate post-hoc testing will be applied if distributional assumptions are not met.

##### Multivariable prediction of vasoplegia

A logistic regression model will be constructed with vasoplegia as the dependent variable. Candidate predictors will include Δrenin (categorised or continuous), ΔBio-ADM, patient characteristics (age, sex, frailty, comorbidities), surgical factors (urgency, procedure type, duration), intra-operative variables (vasopressor exposure, fluid balance, epidural use), and baseline medication use (including RAAS inhibitors and beta blockers). Variables will undergo univariable screening prior to multivariable modelling. Model performance will be assessed through discrimination (C-statistic), calibration, and internal validation using bootstrapping.

##### Renin and acute kidney injury (AKI)

Δrenin will be compared between patients who do and do not develop AKI. ROC analysis will evaluate its predictive performance. Longitudinal differences in renin trajectories between AKI groups will be assessed using mixed-effects modelling.

##### Renin and organ dysfunction

Associations between Δrenin and the composite organ dysfunction outcome will be examined using appropriate univariable analyses.

##### Vasopressor exposure

Correlations between Δrenin and markers of vasopressor burden (peak dose, duration, cumulative exposure) will be assessed using parametric or non-parametric correlation methods as appropriate.

##### Endothelial biomarkers

Baseline concentrations and peri-operative changes in Bio-ADM and syndecan-1 will be compared across outcome groups. Their predictive performance will be evaluated using ROC analysis and incorporated into multivariable models where appropriate.

##### Proenkephalin and AKI

Early post operative proenkephalin levels will be compared between patients with and without subsequent AKI. Predictive performance will be assessed using ROC analysis, and incremental value beyond creatinine change will be explored.

##### Clinical complications

The incidence of cardiovascular complications, sepsis, and need for mechanical ventilation will be compared between vasoplegia groups using appropriate categorical tests.

### Exploratory/Tertiary Analyses

#### Immune Biomarkers

cDPP3:

- Compare pre-operative and 4h post operative levels between outcome groups (vasoplegia, AKI, prolonged ICU stay, mortality)
- Assess persistent elevation (lack of decrease from 4h post-op to POD1) and association with mortality
- Dichotomize by clinically relevant threshold (>30 ng/mL) and calculate odds ratio for mortality using Fisher’s exact test

DPP4:

- Assess whether patients on DPP4 inhibitors (gliptins) have lower incidence of vasoplegia or AKI
- Fisher’s exact test comparing outcomes; calculate odds ratio with 95% CI

suPAR:

- Compare pre-operative and 4h post operative levels between outcome groups
- ROC analysis for outcome prediction
- Assess potential as point-of-care risk stratification tool

Subphenotyping by Angiopoietins:

- Calculate Angiopoietin-1/Angiopoietin-2 ratio
- Categorize patients into high vs. low Ang-1/Ang-2 ratio (median split or data-driven cut-off)
- Compare clinical outcomes between subphenotypes
- K-means clustering or latent class analysis to identify distinct endothelial dysfunction patterns

Inflammatory Subphenotypes:

- Latent class analysis using cytokine profiles (sTNFR-1, others if measured)
- Compare outcomes across identified inflammatory subphenotypes
- Exploratory hypothesis generation regarding mechanistic pathways

Interaction Effects:

Explore potential effect modification::

- Emergency vs. elective surgery
- Presence of peritonitis
- Baseline RAAS inhibitor use
- Baseline renal function (eGFR categories)
- Epidural use

Statistical testing: Include interaction terms in regression models; likelihood ratio test for significance.

Subgroup Analyses:

Pre-specified subgroup analyses for primary outcome (Δrenin and vasoplegia):

- Surgical urgency: Elective vs. emergency
- Surgical indication:Infection/peritonitis vs. other indications
- RAAS medication use: ACE inhibitor/ARB users vs. non-users (with attention to perioperative continuation vs. omission)
- Baseline renal function: eGFR ≥60 vs. <60 mL/min/1.73m²
- Epidural use: Present vs. absent
- Surgery type: Upper GI/hepatobiliary vs. gynecologic vs. urologic vs. colorectal

Analysis Approach:

- Descriptive comparison of effect estimates across subgroups
- Formal interaction testing using regression models
- Forest plots displaying subgroup-specific effect estimates with 95% CIs
- No multiplicity adjustment; interpreted as exploratory

### Software

All statistical analyses will be performed using: R version 4.0 or higher (R Foundation for Statistical Computing, Vienna, Austria), Key packages: tidyverse (data manipulation), lme4 (mixed models), pROC (ROC analysis), mice (multiple imputation), rms (regression modeling strategies), ggplot2 (visualization). GraphPad Prism 9 for figure generation (if needed). Statistical code will be version-controlled and archived to ensure reproducibility.

### Reporting

Results will be reported in accordance with STROBE (Strengthening the Reporting of Observational Studies in Epidemiology) guidelines [16]. All pre-specified analyses will be reported regardless of statistical significance. Deviations from this statistical analysis plan will be documented and justified in the final publication. Effect estimates will be reported with 95% confidence intervals and exact p-values (avoiding “p<0.05”). Clinical interpretation will emphasize effect sizes and confidence intervals over p-values alone.

## Data Availability

All data produced in the present study are available upon reasonable request to the authors

## Abbreviations/Glossary

ACE: Angiotensin Converting Enzyme
ADM: Adrenomedullin
AKI: Acute kidney injury
ASA: American Surgical Association
Bio-ADM: Bioactive adrenomedullin
ESICM: European Society of Intensive Care Medicine
KDIGO: Kidney Disease Improving Global Outcomes
MAR: Missing at Random
MCAR: Missing Completely at Random
MICE: Multiple imputation by chained equations
MNAR: Missing Not at Random (MNAR)
Δrenin - delta renin: Change in renin between pre-operative and initial post operative levels
PenKID: Proenkephalin
POD1: Post-operative day 1
POD2: Post operative day 2
RAAS: Renin Angiotensin Aldosterone System

## Appendices

### Case Report Form

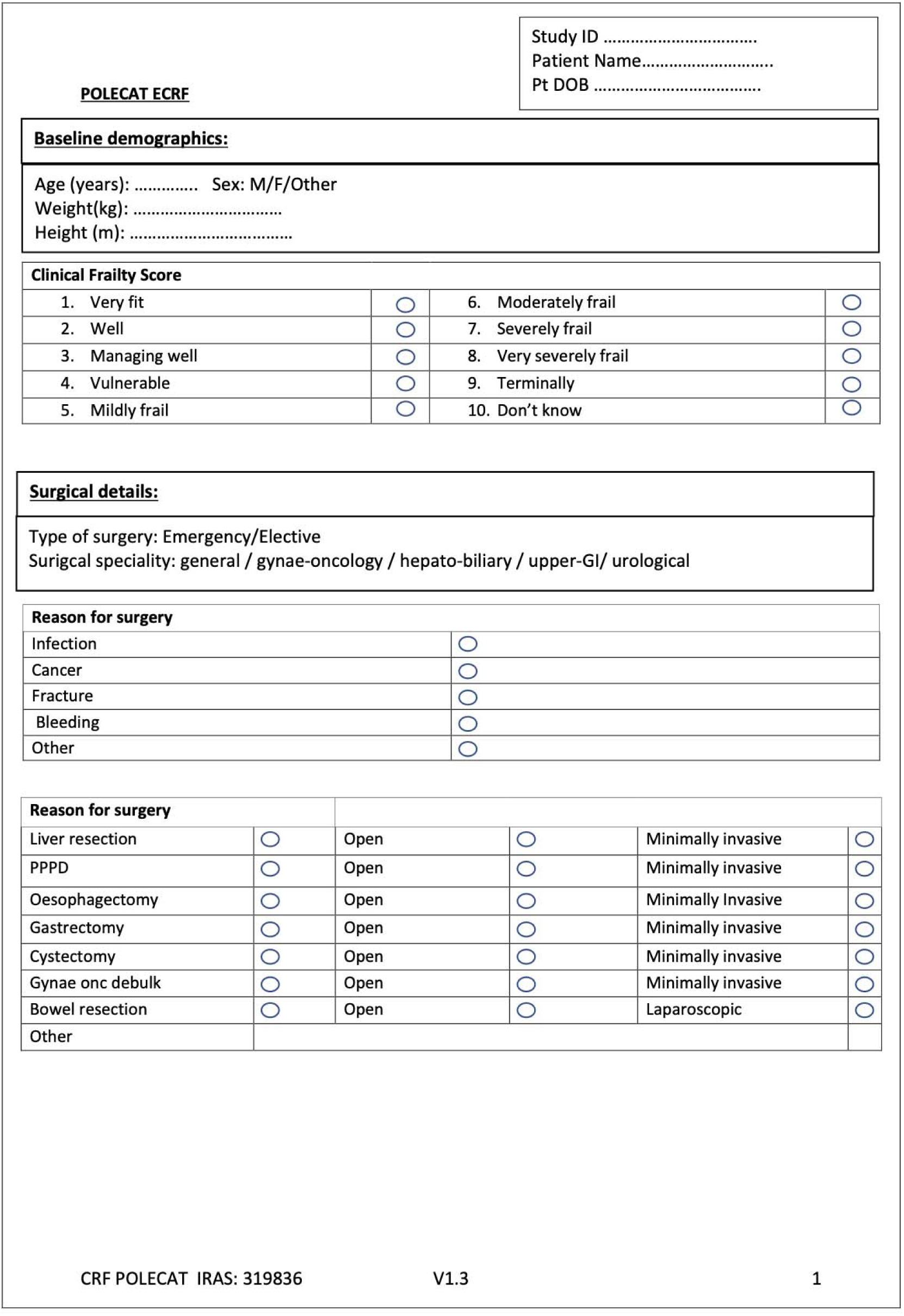
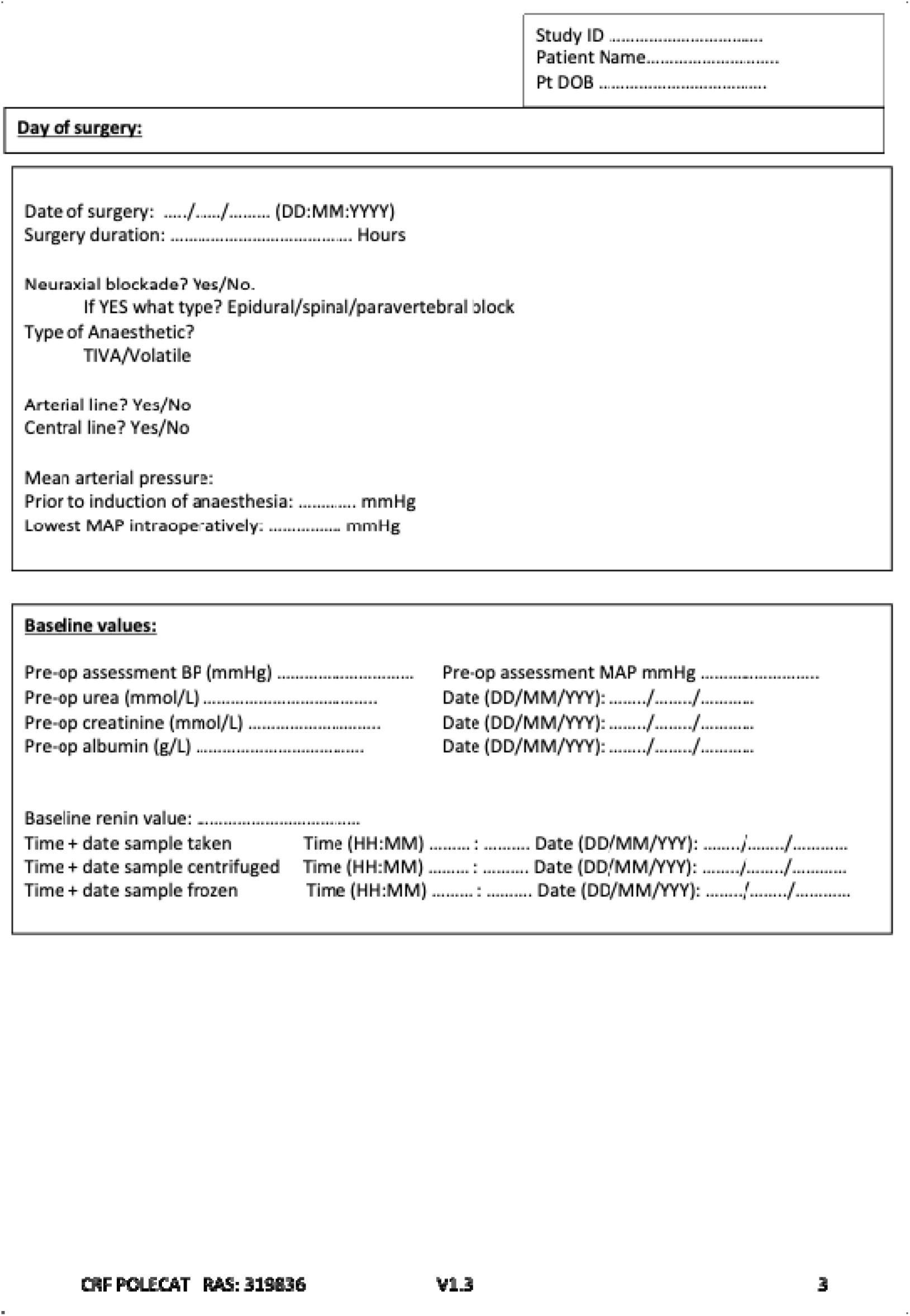
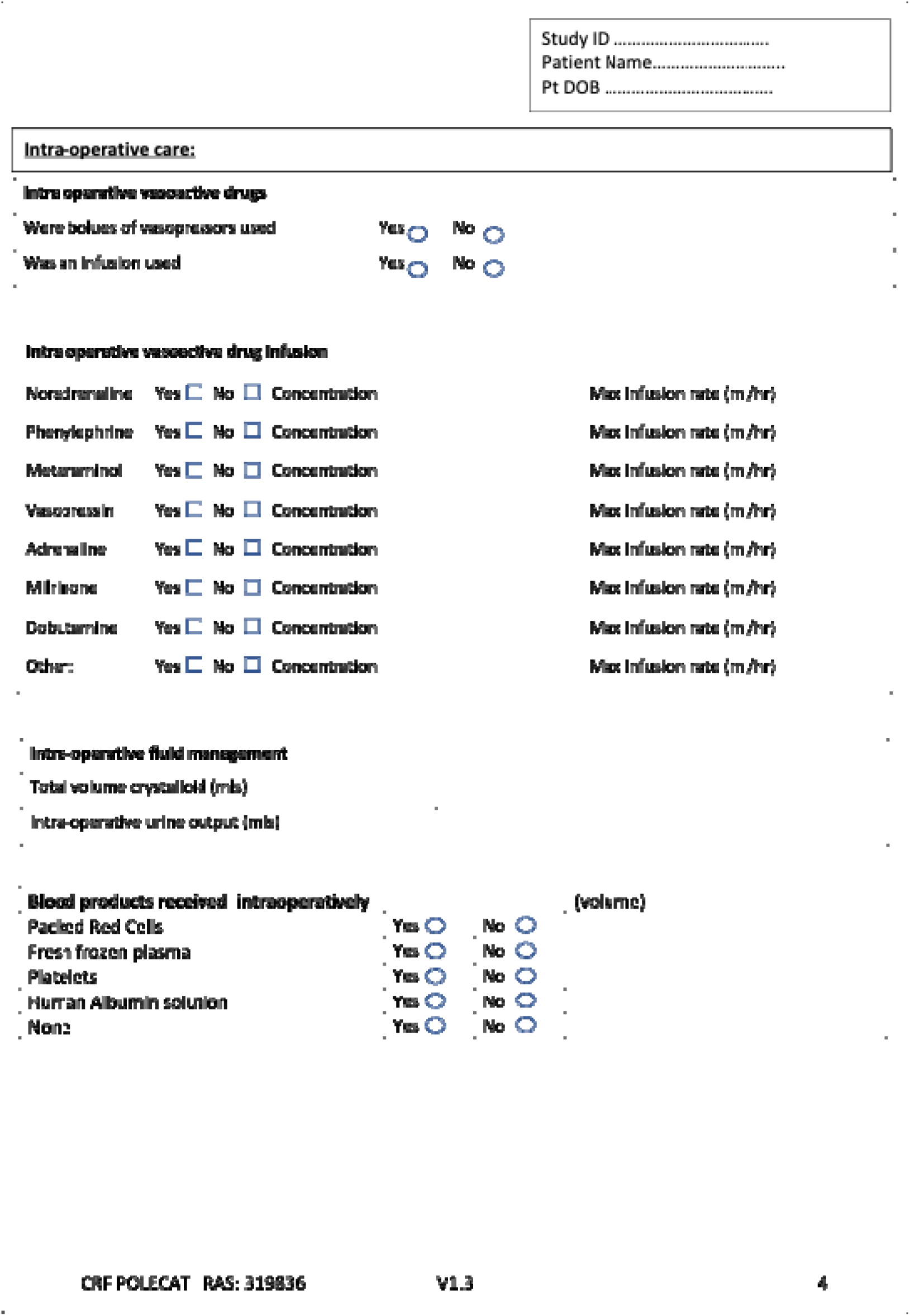
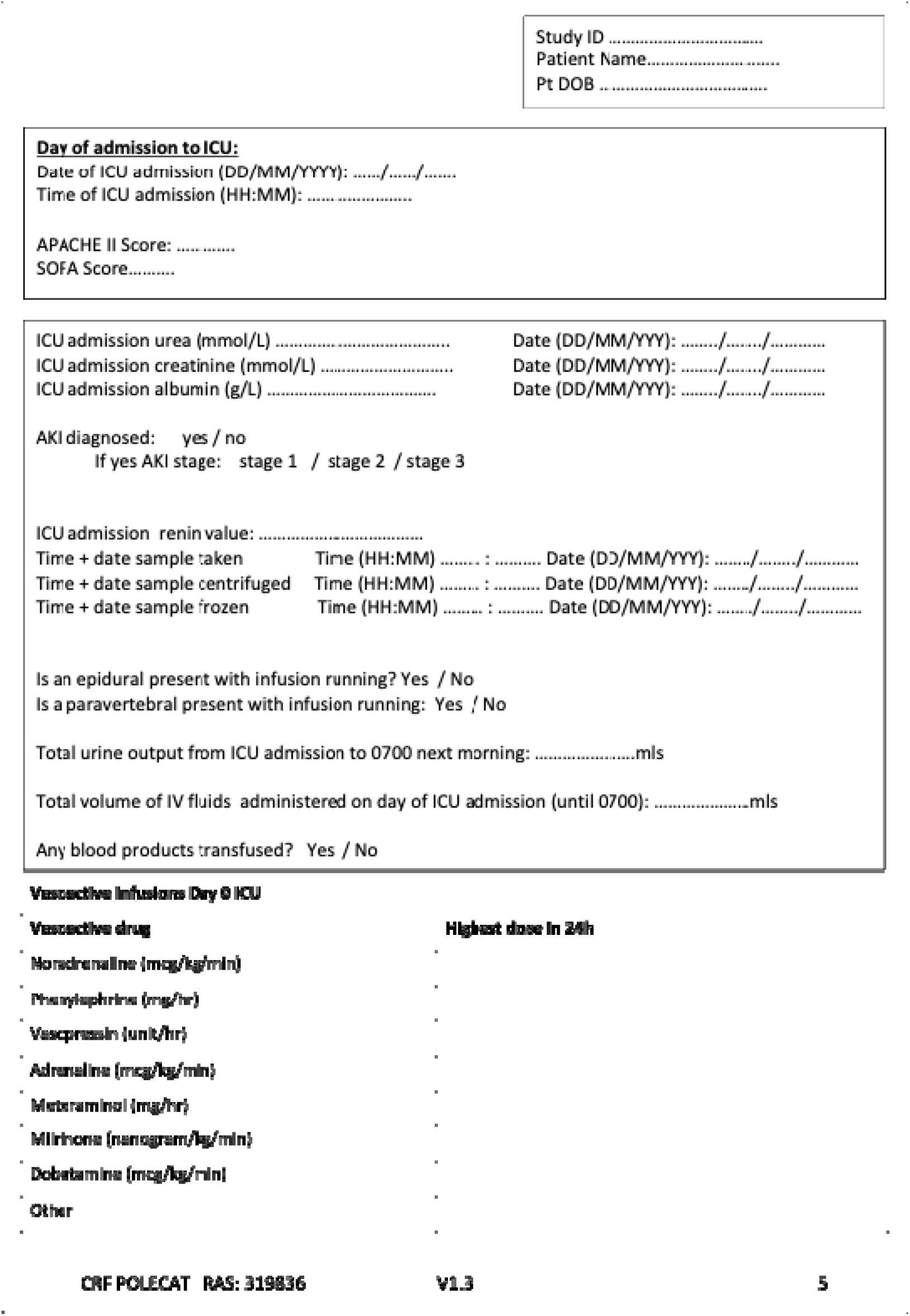
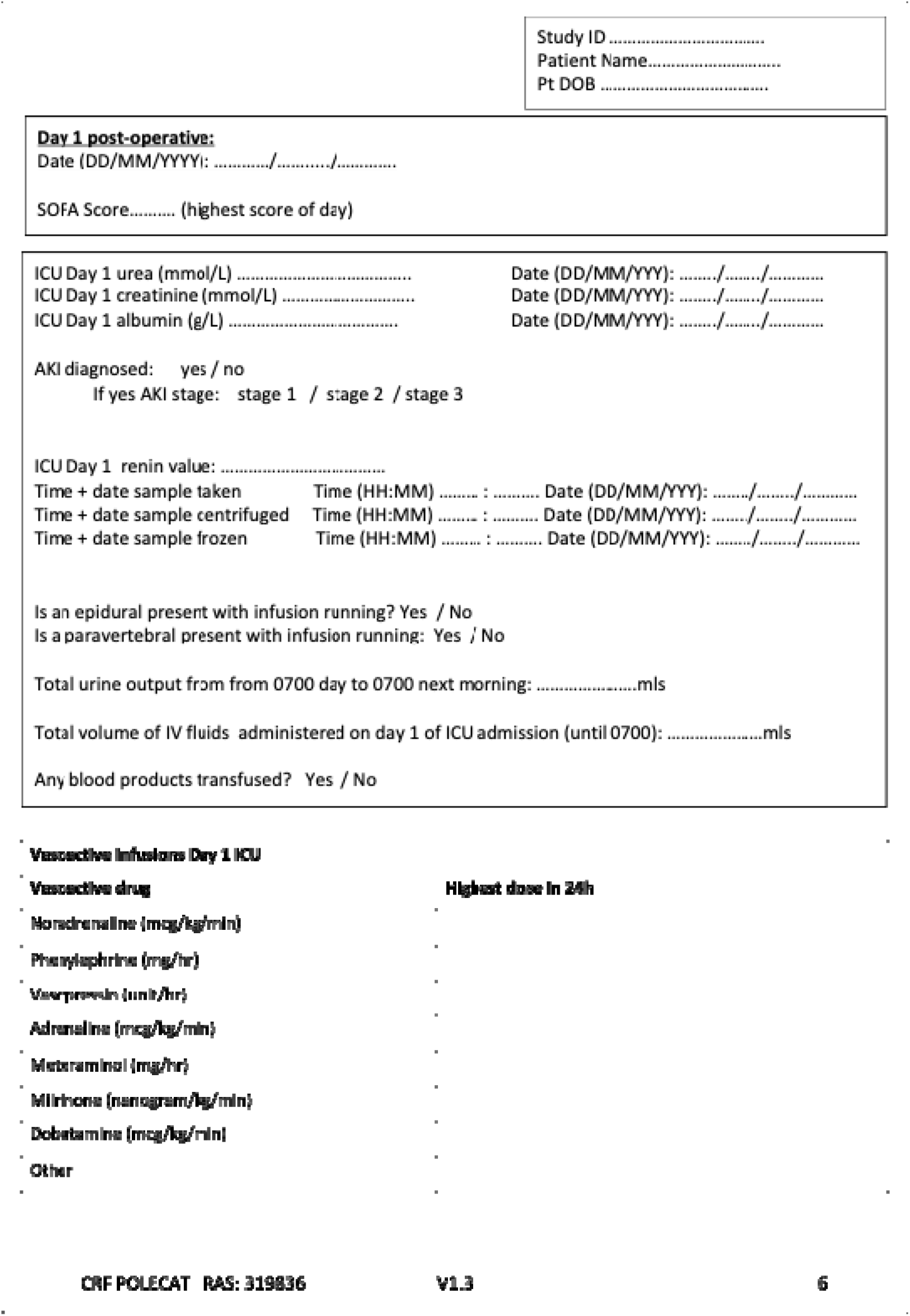
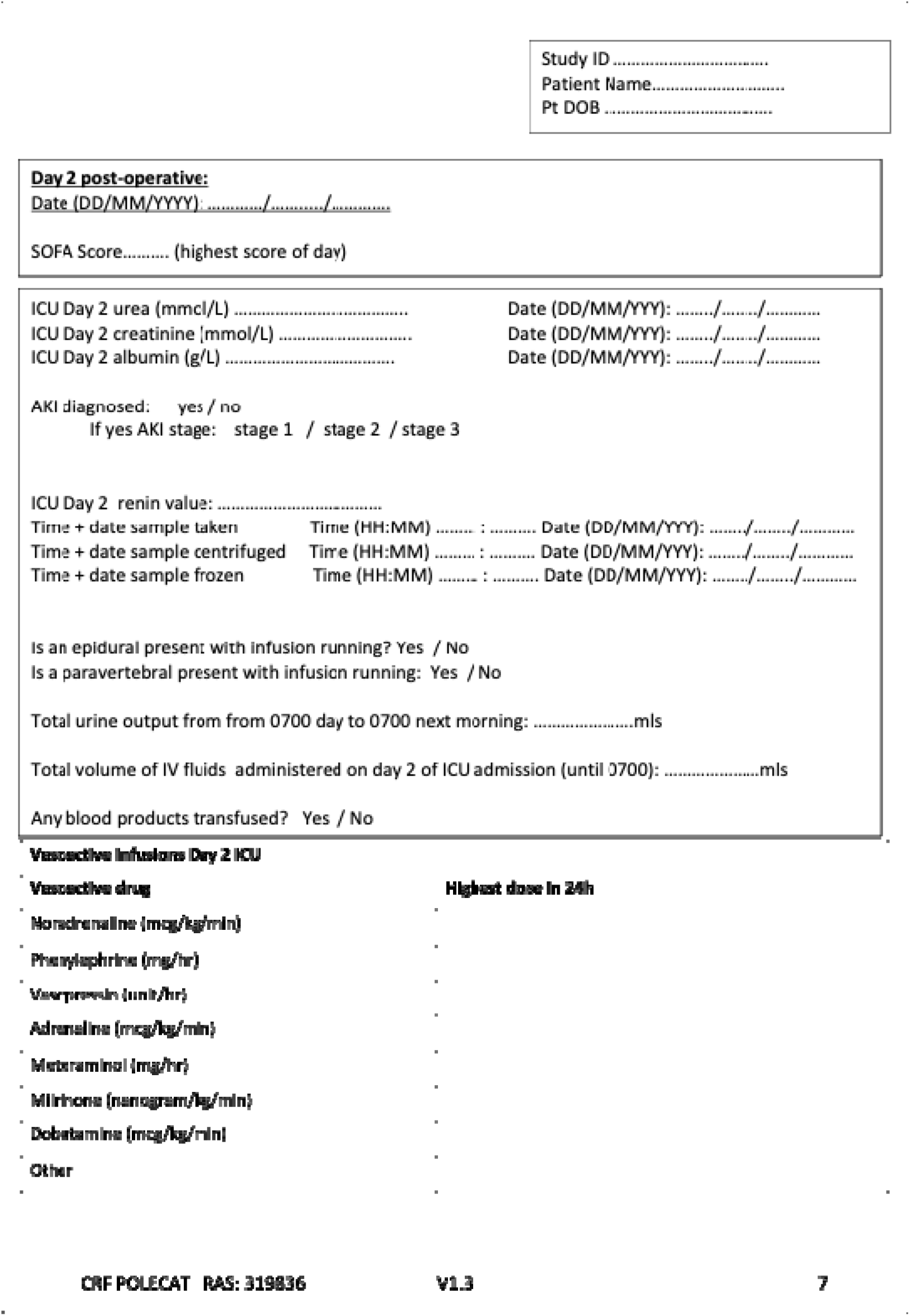
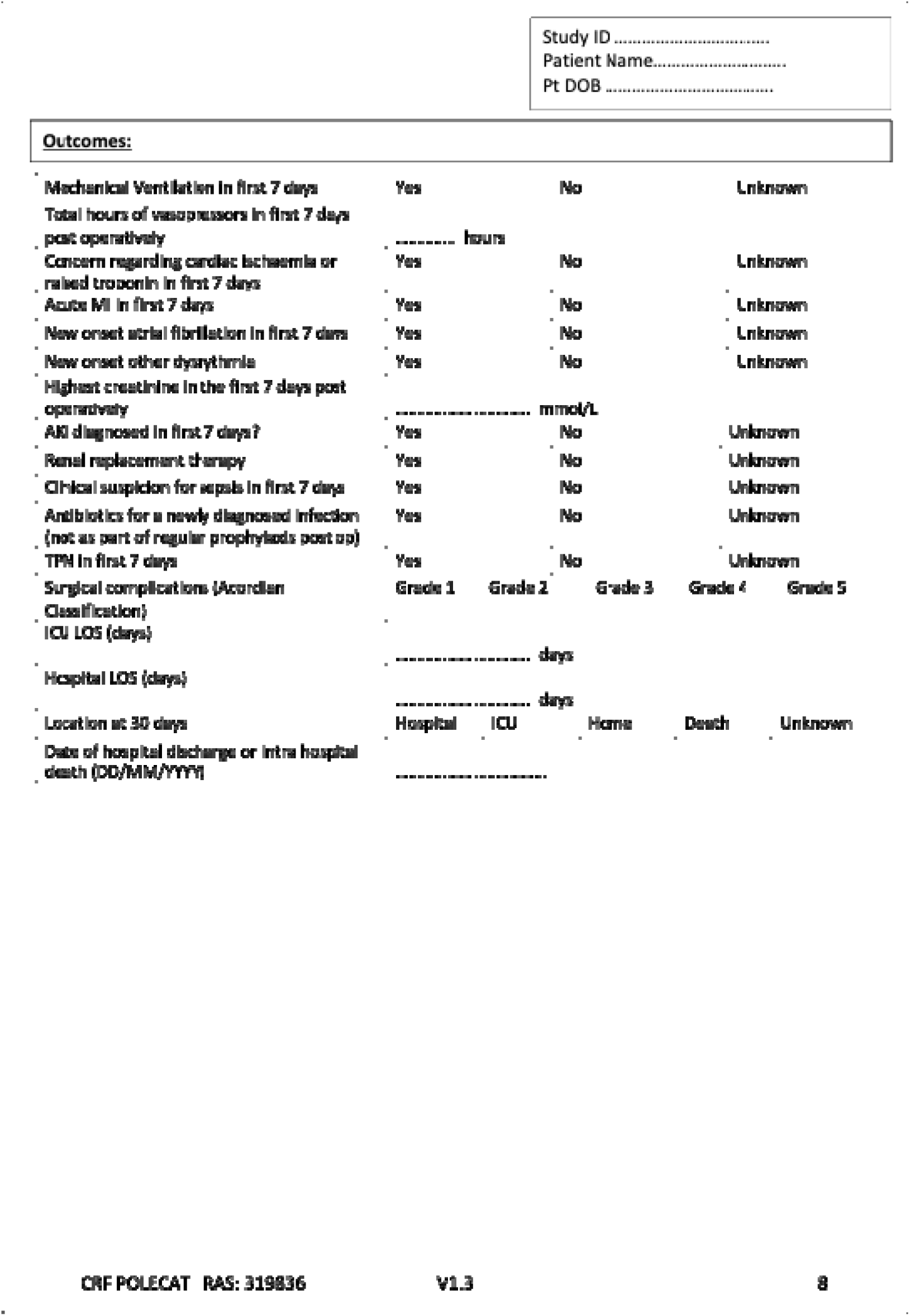

### REC Approval Letter

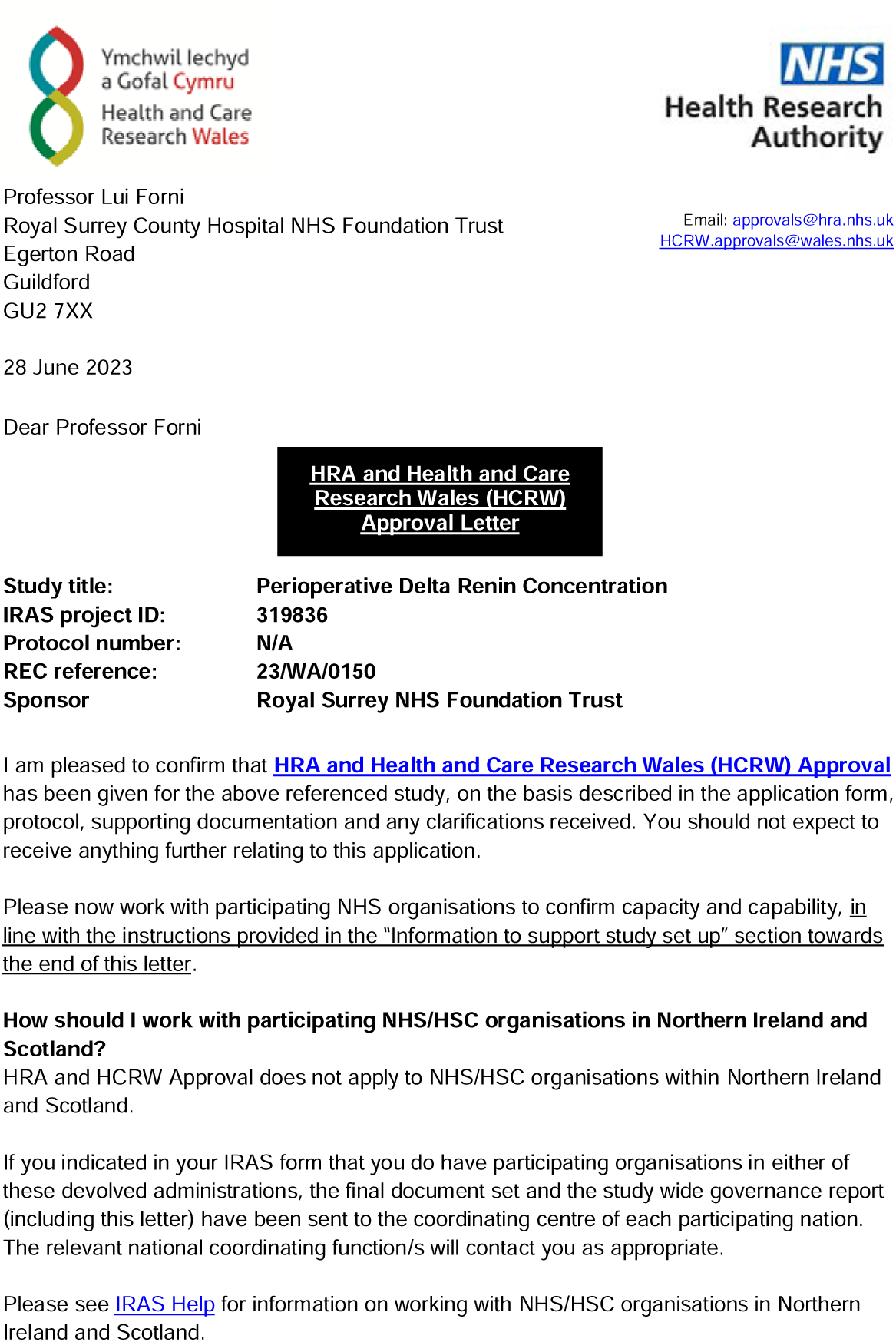
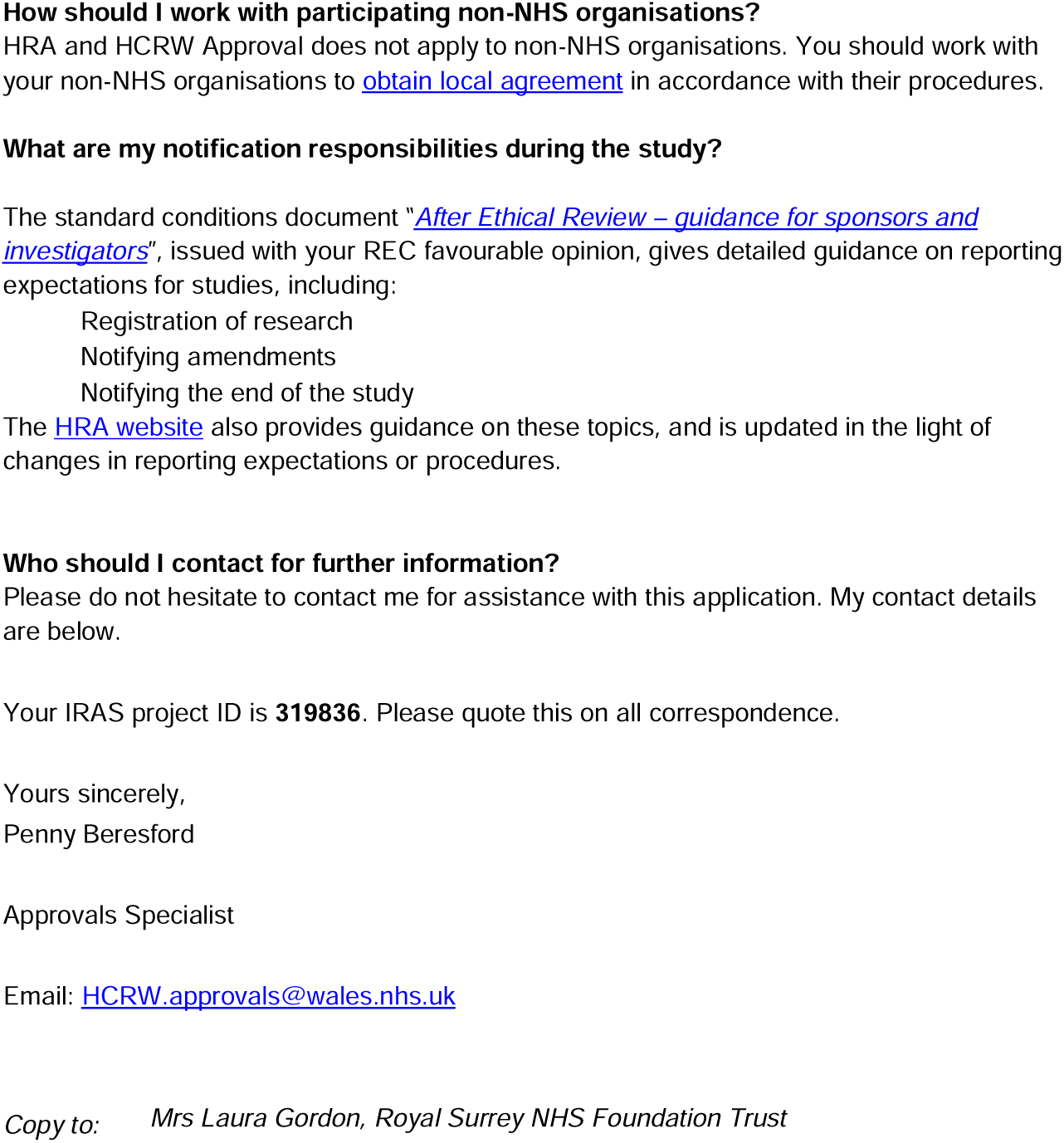

## Notes

### Competing Interest Statement

AZ has received consulting fees from Astute-Biomerieux, Baxter, Bayer,
Novartis, Guard Therapeutics, AM Pharma Paion, Viatris, Renibus, Alexion, Fresenius research funding from Astute-Biomerieux, Fresenius, Baxter, and speakers fees from Astute-Biomerieux Fresenius Baxter.

### Funding Statement

This study was funded by ESICM Levi-Montalcini Biomedical Sciences Award

### Author Declarations

HRA and Health and Care Research Wales IRAS 319836 REC reference 23/WA/0150

